# Obesity and COVID-19: The role of visceral adipose tissue

**DOI:** 10.1101/2020.05.14.20101998

**Authors:** Antonia Petersen, Keno Bressem, Jakob Albrecht, Hans-Martin Thieß, Janis Vahldiek, Bernd Hamm, Marcus R. Makowski, Alexandra Niehues, Stefan Niehues, Lisa C. Adams

**Affiliations:** Charité – Universitätsmedizin Berlin, Campus Benjamin Franklin – Department of Radiology, Hindenburgdamm 30, 12203 Berlin, Germany; Technical University of Munich, School of Medicine, Department of Diagnostic and Interventional Radiology, 81675 Munich, Germany; Charité – Universitätsmedizin Berlin, Campus Charité Mitte – Department of Radiology, Charitéplatz 1, 10117 Berlin, Germany; Berlin Institute of Health (BIH), 10178 Berlin, Germany

**Keywords:** COVID-19, SARS-CoV-2, obesity, quantification of adipose tissue, visceral adipose tissue

## Abstract

**INTRODUCTION:** During the unprecedented health crisis of the COVID-19 pandemic it was suggested that obesity might aggravate severe acute respiratory syndrome coronavirus-2 (SARS-CoV-2). Therefore, this study aims to investigate the association between Compute Tomography (CT)-based measurements of visceral and subcutaneous fat as measures of obesity and COVID-19 severity.

**METHODS:** 30 patients with laboratory-confirmed COVID-19 and a mean age of 65.59 ± 13.06 years from a level one medical center in Berlin, Germany, were retrospectively analyzed and included in the present analysis. SARS-CoV-2 was confirmed by polymerase chain reaction from throat swaps or deep nasal swabs on the day of admission. Severe clinical courses of COVID-19 were defined by hospitalization in intensive care unit (ICU) and invasive mechanical ventilation. All patients received low-dose chest CT-based fat measurements at the level of the first lumbar vertebra.

**RESULTS:** An increase in visceral fat area (VFA) by one square decimeter was associated with a 22.53-fold increased risk for ICU treatment and a 16.11-fold increased risk for mechanical ventilation (adjusted for age and sex). For upper abdominal circumference, each additional centimeter of circumference showed a 1.13-fold increased risk for ICU treatment and a 1.25-fold increased risk for mechanical ventilation. There was no significant correlation of area (SFA) or body mass index (BMI) with severe clinical courses of COVID-19.

**CONCLUSIONS:** Our results suggest that visceral adipose tissue and upper abdominal circumference specifically increasing the risk of COVID-19 severity. CT-based quantification of visceral adipose tissue and upper abdominal circumference in routinely acquired chest CTs may therefore be a simple tool for risk assessment in SARS-CoV-2-patients.

## Introduction

In the context of the present unprecedented health crisis due to the COVID-19 pandemic, it was suggested, that obesity might also aggravate severe acute respiratory syndrome coronavirus-2 (SARS-CoV-2) (1, 2). Previous research only suggested obesity in general, based on the individual body mass index (BMI), as a risk factor for severe COVID-19, without making a distinction between subcutaneous and visceral fat. However, next to overall obesity, body fat distribution is an important risk factor for cardio-metabolic outcomes (1, 2). Especially visceral fat accumulation is often associated with metabolic syndrome, conferring an increased risk of cardiovascular disease and type 2 diabetes with a subsequently enhanced morbidity (1, 2). Furthermore, while BMI is currently the most frequently used indicator for assessing overweight/obesity in adults with a high specificity, it has an inherent low sensitivity to identify excess fat mass and cannot distinguish between visceral and subcutaneous fat (3). Here, computed tomography (CT) with non-invasive post-processing applications allows for differentiation between and quantification of visceral and subcutaneous fat tissue and thus for a reliable assessment of body fat distribution (4).

Therefore, the aim of this study was to quantify the subcutaneous and visceral abdominal fat of patients with a SARS-CoV-2 infection based on routinely acquired low dose chest CTs and to correlate these results with the rate of hospitalization in ICU and invasive mechanical ventilation as severe clinical courses.

## Material and Methods

This retrospective study was prior approved by the institutional review board and carried out between March 2020 and April 2020, including all patients with SARS-CoV-2, who presented at the emergency department of our institution or were transferred from other sites. Presence of SARS-CoV-2 was confirmed by polymerase chain reaction from throat swaps or deep nasal swabs on the day of admission.

### 2.1. CT imaging protocol

For CT imaging, two different types of scanners were used: Three Canon Aquilion PRIME (CAP) scanners and a GE Lightspeed VCT (GEL) scanner. A low dose chest CT was performed for diagnostic purposes or to evaluate the patient’s current condition on the day of admission. The imaging protocol was based on the following imaging parameters: Highest rotation time available CAP: 0.27 s, GEL 0.35 s, 100 kV, automatic tube modulation between 10 and 100 mA with a noise index of 27 (CAP) and 39 (GEL). For CAP, the collimation used was 80 × 0.5 mm, for GEL, it was 64 × 0.625 mm. Pitch factors were 1.388 (CAP) and 1.375 (GEL). Non-enhanced scans were performed in deep inspiration with caudocranial spiral acquisition and supine positioning of the patient. We reconstructed 0.5 mm (CAP) and 0.625 mm (GEL) axial slices in soft tissue and lung kernel reconstruction (CAP: Fc01 and Fc85, GEL: “standard” and “lung”), using iterative reconstruction technology (CAP: AIDR3D level “moderate”, GEL: ASIR 50%).

### 2.2. Data acquisition

CT fat measurement was performed based on chest CTs using Vital’s Vitrea^™^ Advanced Visualization applications (Version 7.0, Canon Medical Systems Cooperation, Otawara, Tochigi, Japan). Visceral fat area (VFA), subcutaneous fat area (SFA) and total fat area (TFA) as well as and visceral fat area/total fat area (VFA/TFA) were quantified at the level of the mid of the first lumbar vertebra on thoracic CT scans acquired in routine clinical imaging for COVID-19 (5, 6). Data was then exported as comma-separated values.

Clinical courses were obtained from conventional chest radiographs, which, at our site, are usually taken on the day of admission to the ICU and/or after initiation of mechanical ventilation. Other patient information such as age, weight or height were extracted from the DICOM headers of the CT scans, if available. The body mass index (BMI) was calculated according to the usual formula: weight (kg) / [height (m)]^2^. All acquired data were stored in tabular form and then exported as comma-separated values for further statistical analysis.

### 2.3. Statistical Analysis

All statistical analysis was performed using the ‘R’ statistical environment (Version 4.0.0), including the “tidyverse” library (7, 8). The different variables showed different value ranges. To facilitate comparison, continuous variables were scaled to a uniform level before analysis. Continuous variables were tested for normal distribution using the Shapiro-Wilk test. If normally distributed, they were expressed as mean ± standard deviation. Otherwise they were presented as median and interquartile range. Continuous and normally distributed variables were compared with one-tailed Student’s t-test, while the Wilcox rank sum test was used in case of non-normal upper abdominal circumference distribution. Categorical variables were expressed as frequencies and percentages and compared using Fisher’s exact test or the chi-squared test as appropriate. A multivariate binomial logistic regression model was used to analyze the relationship between severe clinical courses and quantitative CT features. 95% confidence intervals for odds ratios were calculated using bootstrapping (1000 iterations). A p-value < 0.05 was considered statistically significant.

## Results

In the present study, we analyzed chest CT images of 30 patients with confirmed SARS-CoV-2 infection. The mean age was 65.6 ± 13.1 years, including 18 males (67.2 ± 17.0 years) and 12 females (71.17 ± 11.8 years). In total, 43 % of the patients had to be transferred to the ICU during treatment (n = 13), with men being affected more often than women (men n = 9, women n = 4, p = 0.47). Out of these 13 ICU patients, 7 required mechanical ventilation (men n = 6, women n = 1, p = 0.19). Please refer to Table 1 for a detailed overview of patient characteristics.

**Table 1.** Patient characteristics.

| Variable | All Patients | ICU | Intubation |
| --- | --- | --- | --- |
| Age | 65.59 ± 13.06 | 63.6 ± 15.11 | 59.34 ± 14.47 |
| Number of patients | 30 | 13 | 7 |
| Total fat area (dm<sup>2</sup>) | 1.51 IQR: 0.76 | 1.9 IQR: 0.84 | 2.37 ± 1.34 |
| Subcutaneous fat area (dm<sup>2</sup>) | 0.62 IQR: 0.48 | 0.87 IQR: 0.57 | 0.73 IQR: 0.6 |
| Visceral fat area (dm<sup>2</sup>) | 0.82 IQR: 0.55 | 1.12 ± 0.53 | 1.24 ± 0.66 |
| Visceral fat area/<br>total fat area | 0.53 ± 0.16 | 0.53 ± 0.17 | 0.56 ± 0.17 |
| Upper abdominal circumference<br>(cm) | 102.54 SD 8.91 | 106.98 ± 8.32 | 109.71 ± 7.62 |
| Visceral fat area/ subcutaneous fat<br>area | 1.38 ± 0.81 | 1.41 ± 0.87 | 1.51 ± 0.74 |
| BMI | 26.37 ± 3.64 | 25.62 ± 1.98 | 24.92 ± 1.34 |

Table 1 provides an overview of descriptive patient characteristics.

### 3.1. Difference between groups

Mean BMI for all patients was 26.4 ± 3.6 (men 25.6 ± 2.4 vs. women 28.3 ± 5.6). Patients who were admitted to the ICU showed a higher BMI (26.8 ± 4.4) compared to patients who did not require ICU treatment (25.6. ± 2.0), but this difference was not significant (p = 0.11).

The median VFA was 0.82 dm^2^ (IQR 0.55 dm^2^). Men tended to have more visceral adipose tissue than women (0.95 dm^2^ vs 0.67 dm^2^), which showed to be a statistically significant difference (p = 0.014). Patients admitted to the ICU had a significantly higher TFA compared to patients treated on the normal ward. When looking at the different fat compartments, it was particularly noticeable that ICU patients showed a significantly larger VFA (1.12 dm^2^, IQR 0.53) when compared to patients treated on the normal ward (0.7 dm^2^, IQR 0.28, p = 0.031). Furthermore, patients requiring mechanical ventilation showed a significantly higher VFA than patients, who could breathe freely (1.24 dm^2^ versus 0.77 dm^2^, p = 0.006) (also refer to Figure 1 for patient case examples). Upper abdominal circumference also significantly differed between patients in ICU (107.0 cm, IQR 8,3) and patients on the normal ward (99.2 cm, IQR 8.0, p = 0.009) and similarly between patients requiring mechanical ventilation (109.7 cm, IQR 7,6) and patients without mechanical ventilation (100.3, IQR 8.2, p = 0.008).

**Figure 1.**
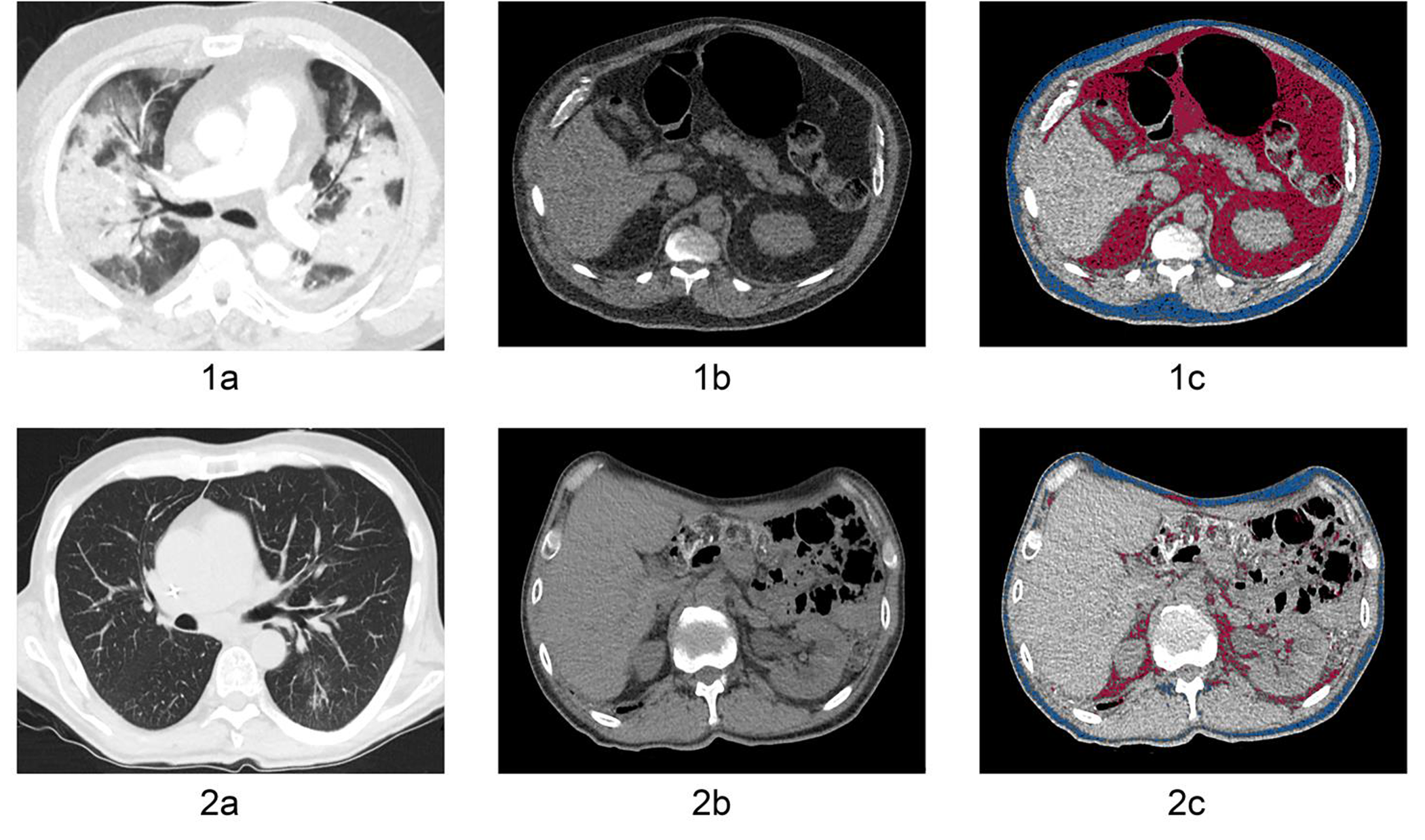
Figure 1 shows two case examples of a lean and an obese patient with COVID-19. 2a-c demonstrate the chest (1a) and abdomen CT (1b) of an obese 66-year-old male patient. 1c shows the CT-based fat quantification of the subcutaneous (blue color) and visceral adipose tissue (red color) with a visceral fat area of 220.4 cm^2^, a subcutaneous fat area of 112.4 cm^2^ and an upper abdominal circumference of 120.4 cm. 2a-c demonstrate the chest (2a) and abdomen CT (2b) of a lean 51-year old male patient. 2c shows the CT-based fat quantification of the subcutaneous and visceral adipose tissue with a visceral fat area of 15.4 cm^2^, subcutaneous fat area of 20.8 cm^2^ and an upper abdominal circumference of 95.9 cm. Of these two cases, the patient with the higher fat content has the more severe pulmonary COVID-19 infection.

SFA and the VFA/TFA ratio showed no statistically significant differences between patients on the normal ward or ICU or patients with or without mechanical ventilation. The differences in the various measurements of fat tissue and BMI between patients are summarized in Table 2.

**Table 2.**
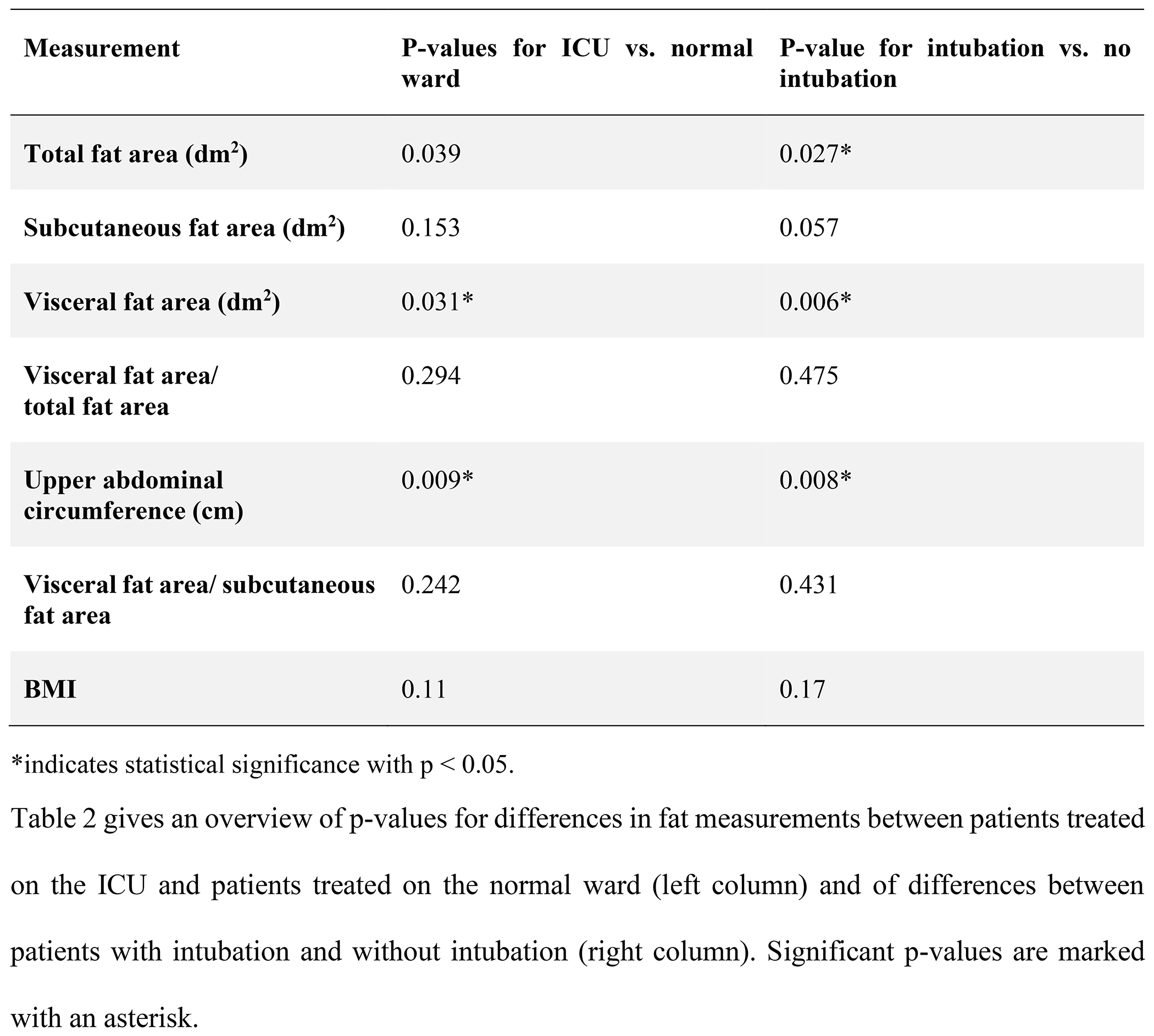
Differences in fat measurements between groups.

### 3.2. Correlation between BMI and fat area measurements

VFA and BMI showed a moderate correlation of 0.56 (95% CI 0.42 – 0.79, p < 0.001). It was noticeable, that the correlation was stronger for men than for women (0.62 vs. 0.53). Upper abdominal circumference and BMI also showed a moderate correlation of 0.61 (95% CI 0.40 – 0.81, p < 0.001). Again, men showed a slightly stronger correlation between BMI and upper abdominal circumference compared to women (0.64 vs. 0.61).

### 3.3. Predictive modeling

As VFA and upper abdominal circumference demonstrated a significant difference between patients, who required treatment on the ICU and/or mechanical ventilation and patients, who did neither require ICU treatment nor mechanical ventilation, a logistic regression analysis was conducted. Two models each were constructed for VFA and upper abdominal circumference, the first unadjusted for any confounder, the second adjusted for age and sex. Table 3 gives an overview of the odds ratios derived from the logistic regression analysis. An increase in VFA by one square decimeter was associated with a 22.53-fold (95% CI 2.01–573.72) increased risk for ICU treatment and a 16.11-fold (95% CI 1.46–642.48) increased risk for mechanical ventilation (adjusted for age and sex). For upper abdominal circumference, each additional centimeter of circumference showed a 1.13-fold (95% CI 1.02–1.3) increased risk for ICU treatment and a 1.25-fold (95% CI 1.05–1.68) increased risk for mechanical ventilation.

**Table 3.** Regression coefficients.

| Unadjusted for age and sex |  |  |  |
| --- | --- | --- | --- |
| Variable | Odds ratio | 95% CI | Outcome |
| Visceral fat area (dm<sup>2</sup>) | 22.1 | 2.17-486.27 | Patient on the ICU |
| Upper abdominal circumference (cm) | 1.13 | 1.03-1.29 | Patient on the ICU |
| Visceral fat area/ total fat area | 1.17 | 0.01-141.35 | Patient on the ICU |
| Visceral fat area (dm<sup>2</sup>) | 13.92 | 1.56-379.04 | Intubation |
| Upper abdominal circumference (cm) | 1.17 | 1.04-1.37 | Intubation |
| Visceral fat area/ total fat area | 5.55 | 0.02-3567.39 | Intubation |
| Adjusted for age and sex |  |  |  |
| Variable | Odds ratio | 95% CI | Outcome |
| Visceral fat area (dm<sup>2</sup>) | 22.53 | 2.01-573.72 | Patient on the ICU |
| Upper abdominal circumference (cm) | 1.13 | 1.02-1.3 | Patient on the ICU |
| Visceral fat area/ total fat area | 0.04 | 0-52.12 | Patient on the ICU |
| Visceral fat area (dm<sup>2</sup>) | 16.11 | 1.46-642.48 | Intubation |
| Upper abdominal circumference (cm) | 1.25 | 1.05-1.68 | Intubation |
| Visceral fat area/ total fat area | 0.01 | 0-120.49 | Intubation |

Table 3 provides the odds ratios derived from multivariate logistic regression analysis, unadjusted and adjusted for age and sex.

## Discussion

In the present study, we quantified subcutaneous and visceral adipose tissue in chest CTs of COVID-19 patients and – apart from the total fat area – identified visceral adipose tissue area and upper abdominal circumference as significant predictors of severe courses of COVID-19, while BMI was not a significant predictor. In our cohort, a severe course was defined by the requirement of ICU treatment and/or mechanical ventilation.

In 2016, the World Health Organization estimated that 1.9 billion adults were overweight and of these, over 650 million were obese (9). Obesity represents a state of low-grade chronic inflammation, which contributes to the development of metabolic diseases, such as dyslipidemia or type 2 diabetes mellitus, and may modify immune responses, which make the immune system more susceptible to infection (8). Obesity-related comorbidities could also represent an additional risk factor in COVID-19 patients, enhancing the risk of severe courses. Given that the epicenters of COVID-19 have moved to Europe and North America, the impact of obesity on COVID-19 may become even more visible, as these two continents have some of the highest prevalence of obesity worldwide, with obesity taking on epidemic proportions (11).

While the previous studies already confirmed general obesity as a risk factor for severe courses of COVID-19, their measurement of obesity was solely based on the measurement of BMI (10–13). However, the utility of BMI in assessing obesity depends on the assumption of a correlation of anthropomorphic measures with more direct measures of obesity, such as visceral and subcutaneous adipose tissue or total body fat (13, 14). Here several studies showed that, for the same BMI, the amount of body fat was influenced by sex and race (15, 16). This also fits with our correlation coefficient of 0,56 for VFA and BMI and the consistent moderate correlation coefficients reported in the literature, ranging from 0,61 to 0,71 after adjustment for age (17–19).

By using CT-based quantification of the visceral and subcutaneous adipose tissue, we chose a more comprehensive approach to measure potential obesity, being able to differentiate between different fat compartments such as subcutaneous and visceral fat. This is essential as body fat distribution was previously established as an independent risk factor for cardio-metabolic outcomes in the general population, next to obesity (2). In the context of our COVID-19 cohort we found, that –apart from TFA – VFA and upper abdominal circumference correlated with ICU treatment and/or mechanical ventilation, while SFA and VFA/TFA did not show any significant correlation with such severe courses of COVID-19. To our knowledge, this is the first study specifically identifying visceral fat and upper abdominal circumference as decisive risk factors for severe courses of COVID-19. This data can be obtained without the need of further imaging based on an automatic analysis of the respective chest CT.

Compared to recent COVID-19 studies from New York and Lille, which only used BMI as a measure of obesity, our ICU patients were similar in terms of age (average age of 65,6 ± 13,1 years) compared to the patients from New York (average age of 67 years, interquartile range 56 – 77), while patients from Lille were younger (average age of 60 years, interquartile range 51 – 70) (17). Regarding median BMI, the Lille centre found that SARS-CoV-2 patients with a BMI ≥ 35 kg/m^2^ required mechanical ventilation significantly more often than SARS-CoV-2 patients with a BMI below 25 kg/m^2^(15). In a centre from New York, especially a BMI ≥ 40 kg/m^2^ was identified to be among the factors most associated with hospitalization of SARS-CoV-2 patients (16). *Zheng et al*. reported patients with metabolic associated fatty liver disease and obesity (BMI > 25 kg/m^2^) to have a 6-fold increased risk for a severe course of SARS-CoV-2 (2). By comparison, we found a more than 23-fold increased risk for ICU treatment and a more than 16-fold increased risk for mechanical ventilation given an increase in VFA by one square decimetre.

The present study also has some limitations, that should be noted: First, it is single-centre design with a relatively small sample size. Second, the study design is retrospective, which can estimate associations, but is incapable of evaluating the temporal sequence. At the end of the observation time many patients were still hospitalized with open final course of the disease. Finally, co-morbidities as well as mortality was not considered and will have to be investigated in larger studies in the future.

### Conclusion

Going beyond the recently established correlation between BMI-based general obesity and severe courses of COVID-19, our results suggest that body fat distribution is also decisive, with visceral adipose tissue and upper abdominal circumference independently increasing the risk for severe courses of COVID-19. Hence, CT-based quantification of visceral adipose tissue might be used as a simple tool for risk assessment in SARS-CoV-2 patients from routinely acquired chest CTs.

## Data Availability

Data available on request due to privacy/ethical restricitons.

## Acknowledgments

MRM is grateful for support from the Deutsche Forschungsgemeinschaft (DFG, SFB 1340/1 2018, 5943/31/41/91). LCA is grateful for her participation in the BIH Charité – Junior Clinician and Clinician Scientist Program funded by the Charité – Universitätsmedizin Berlin and the Berlin Institute of Health.

